# From Tool to Teammate: A Randomized Controlled Trial of Clinician-AI Collaborative Workflows for Diagnosis

**DOI:** 10.1101/2025.06.07.25329176

**Authors:** Selin S. Everett, Bryan J. Bunning, Priyank Jain, Ivan Lopez, Anup Agarwal, Manisha Desai, Robert Gallo, Ethan Goh, Vinay B. Kadiyala, Zahir Kanjee, Jacob M. Koshy, Andrew Olson, Adam Rodman, Kevin Schulman, Eric Strong, Jonathan H. Chen, Eric Horvitz

**Affiliations:** Stanford University School of Medicine, Stanford, CA, USA; Stanford Center for Biomedical Informatics Research, Stanford University, Stanford, CA, USA; Stanford Department of Biomedical Data Science, Stanford, CA, USA; Stanford Clinical Excellence Research Center, Stanford University, Stanford, CA, USA; Stanford Quantitative Sciences Unit, Stanford, CA, USA; Harvard Medical School, Boston, MA, USA; Department of Medicine, Cambridge Health Alliance, Cambridge, MA, USA; University of Minnesota Medical School, Minneapolis, MN, USA; Center for Innovation to Implementation, VA Palo Alto Health Care System, Palo Alto, CA, USA; Department of Medicine, Beth Israel Deaconess Medical Center, Boston, MA, USA; Operations, Information and Technology, Stanford University Graduate School of Business, Palo Alto, CA, USA; Division of Hospital Medicine, Stanford University, Stanford, CA, USA; Office of the Chief Scientific Officer, Microsoft, Redmond, WA, USA; Stanford Institute for Human-Centered Artificial Intelligence, Stanford University, Stanford, CA, USA

## Abstract

Early studies of large language models (LLMs) in clinical settings have largely treated artificial intelligence (AI) as a tool rather than an active collaborator. As LLMs now demonstrate expert-level diagnostic performance, the focus shifts from whether AI can offer valuable suggestions to how it can be effectively integrated into physicians’ diagnostic workflows. We conducted a randomized controlled trial (n=70 clinicians) to evaluate the value of employing a custom GPT system designed to engage collaboratively with clinicians on diagnostic reasoning challenges. The collaborative design began with independent diagnostic assessments from both the clinician and the AI. These were then combined in an AI-generated synthesis that integrated the two perspectives, highlighting points of agreement and disagreement and offering commentary on each. We evaluated two workflow variants: one where the AI provided an initial opinion (AI-first), and another where it followed the clinician’s assessment (AI-second). Clinicians using either collaborative workflow outperformed those using traditional tools, achieving average accuracies of 85% (AI-first) and 82% (AI-second), compared to 75% with traditional resources (p < 0.0004 and p < 0.00001; mean differences = 9.8% and 6.8%; 95% CI = 4.6%–15% and 4.0%–9.6%). Performance did not differ significantly between workflows or from the AI-alone score of 90%. These results underscore the value of collaborative AI systems that complement clinician expertise and foster effective coordination between human and machine reasoning in diagnostic decision-making.

## Introduction

The use of large language models (LLMs) to support physicians’ diagnostic reasoning is in its infancy. Evaluations of GPT-4 on medical challenge problems have demonstrated that LLMs can reach near-expert performance on standardized competency exams such as the United States Medical Licensing Examination (USMLE), fueling growing interest and debate about their potential readiness for real-world clinical use.^1,2^

This study was motivated by unexpected findings in Goh and Gallo et al. that showed physicians using an LLM perform significantly worse than the LLM alone.^3,4^ Insufficient skill and experience with prompting LLMs was discussed as an explanation for the poor AI-assisted human performance. The authors highlighted that, “development in human-computer interactions is needed to realize the potential of AI in clinical decision support.”^3^ We take that charge as the starting point of our investigation.

Inspired by prior research on principles, mechanisms, and designs for human-AI interaction^5–8^, we created a custom GPT-4, guided by a system prompt to support collaboration on diagnostic reasoning. We conducted a randomized controlled trial to gather evidence aimed at informing designs and practices for employing AI models in diagnostic decision-making by evaluating how well participants scored on answering structured reflection questions around clinical diagnostic cases.

This study evaluated whether a collaborative clinician-AI workflow, where the clinician and AI independently assess a case, followed by an AI-generated synthesis and critique, can enhance diagnostic reasoning on complex cases. We also examined two sequencing strategies: using AI as a first opinion versus AI as a second opinion after the clinician’s initial assessment. In addition, we explored how this interactive design influenced clinicians’ engagement with the AI and their attitudes toward its role in diagnostic decision making.

## Methods

We customized a version of GPT-4 (using the GPTs framework) with a system prompt designed to facilitate collaboration between clinicians and the AI in diagnostic reasoning. The prompt instructed the model to act as a diagnostic collaborator: generating its own differential diagnosis, then comparing it with the clinician’s assessment. It then guided the model to produce a synthesis that integrated both perspectives, highlighting areas of agreement and disagreement, offering critiques of each, and inviting further discussion (see **Figure 1**). Details on the system prompt are provided in **Supplementary Figure 1**.

**Figure 1.**
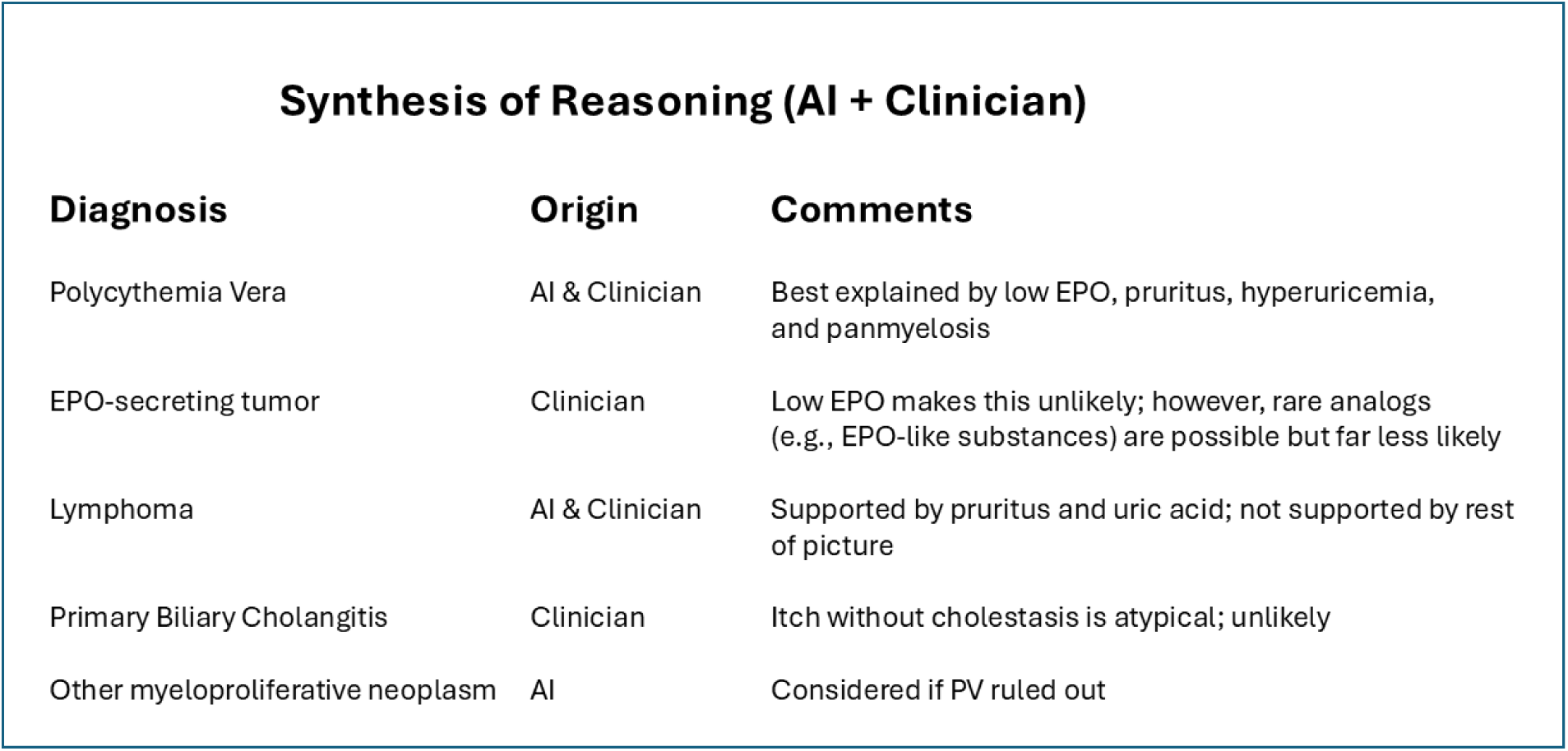
Sample display of synthesis of AI and clinician input with critiques.

We evaluated clinician-AI collaboration in two diagnostic workflows: *AI-as-first-opinion*, where the clinician sees the AI’s output before offering their own, and *AI-as-second-opinion*, where the clinician first provides their answers before consulting the AI. The synthesis view was incorporated into the AI-as-second-opinion workflow immediately following the AI’s recommendations, and was made optionally available to clinicians in the AI-as-first-opinion arm.

We enrolled participants from December 16 2024 to January 24 2025. Inclusion criterion was being a U.S.-licensed internal medicine or family medicine physician. Exclusion criterion was participation in any previous study using the clinical vignettes utilized in our study. We recruited attending and resident physicians via our networks and email lists at Stanford University, Beth Israel Deaconess Medical Center, Vanderbilt University Medical Center, New York-Presbyterian Hospital, and Cambridge Health Alliance. Resident participants were offered $100, and attending participants were offered $199 for completing a one-hour session. Participants joined remotely in small groups (≤ 5). The same study team member (S.E.) facilitated each session, and randomization occurred at the session level. Participant flow is illustrated in **Figure 2**.

**Figure 2.**
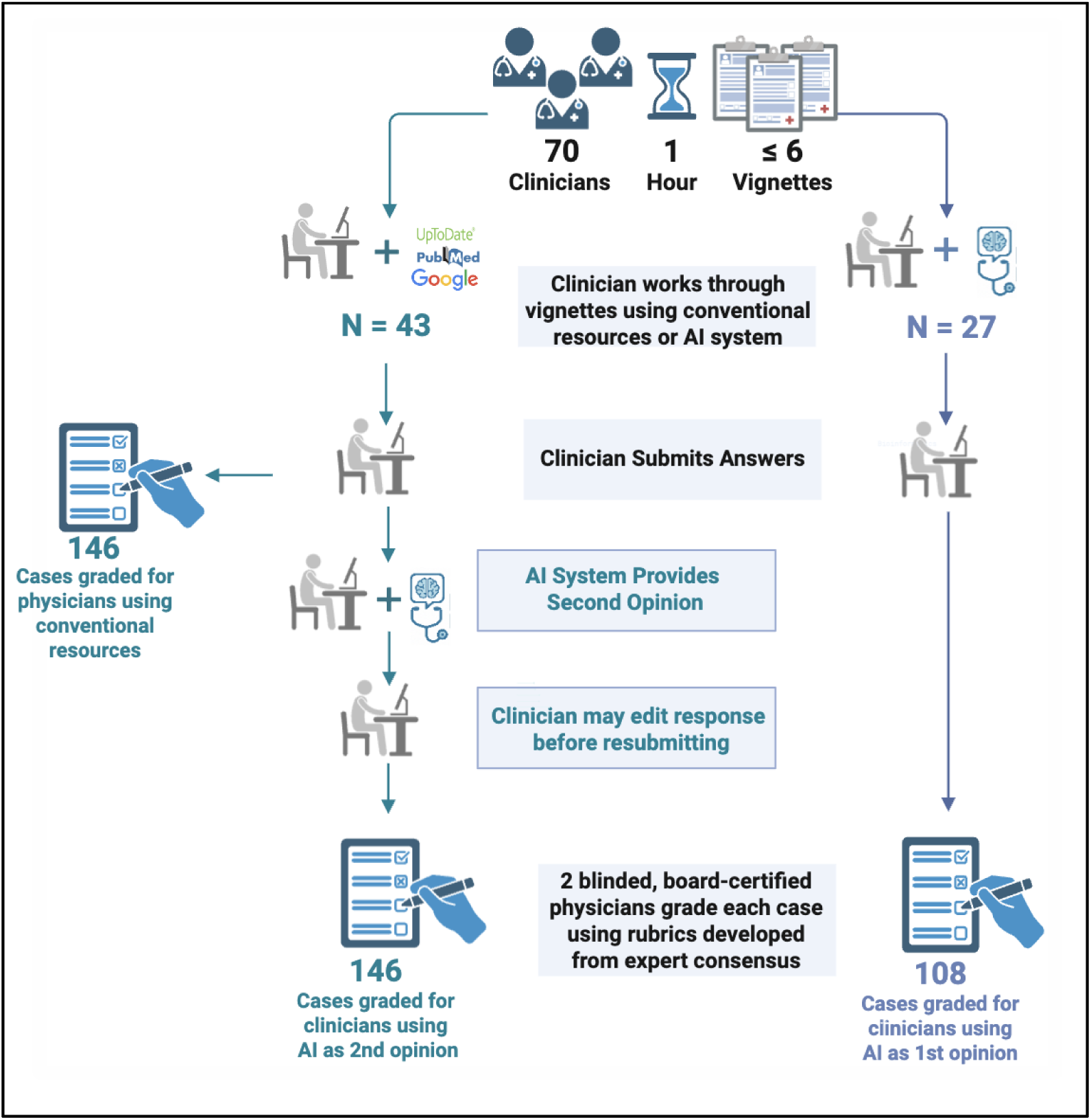
Study workflow.

Participants were encouraged to critically evaluate, challenge, or refine the AI model’s diagnostic conclusions as needed and were reminded that the system outputs could contain errors or gaps. Participants were instructed to start a new AI dialogue for each of the diagnostic reasoning cases to clear the context under consideration by the LLM. Participants completed as many cases as possible in the hour, with instruction to prioritize quality over quantity.

The study was conducted via a secure survey platform (Qualtrics). At the start of each session, the facilitator provided a brief, live demonstration of the case workflow to clearly show that the vignette was to be entered via a copy-paste operation into the AI. For the facilitator’s full script used in each session, please refer to the standard operating procedures included in the **Supplementary Methods 1**. Support from the facilitator was available throughout the session. All participants evaluated a set of up to six clinical vignettes, with their order randomized. The evaluation format, response structure, and case vignettes were identical to the approach presented previously.^3^ Unfinished cases in the study period were excluded. A case was considered unfinished if (1) it was the final vignette in a session, and (2) the participant spent less than 30 seconds reviewing information.

The vignettes are based on real patients. The content was deidentified and included information about history, physical examination, and laboratory test results (**Supplementary Figure 3).** The cases have not been publicly released and therefore are excluded from the training data of the LLM.

### Two workflows

A single custom GPT was designed to support two distinct workflows, depending on the arm a clinician was randomized to. Of note, the system was intentionally designed to broaden clinician reasoning, offering five differential diagnoses rather than three, as requested in the instructions, and to suggest seven management steps instead of three. The model was instructed to rank the differential diagnoses by likelihood. Case response structure is shown in **Supplementary Figure 2**.

For the *AI-as-first-opinion* arm, physicians began by inputting the full case and had the opportunity to use the AI system from the outset to generate ideas, interpret information, and construct their responses. In the *AI-as-second-opinion,* physicians initially worked independently, forming their own assessments with access to conventional (non-AI/LLM) resources such as UpToDate, PubMed, or Google. Participants were instructed to paste the vignette along with their initial answers into the system to initiate a collaborative interaction. After reviewing the AI second opinion, clinicians had to revise or retain their original responses. After completing the initial stage of the structured workflow and viewing the integrated summary, participants were invited by the system to engage in free-form dialogue before submitting their final answers.

#### Baselines for clinicians and AI

To establish a baseline for clinician performance, we scored the initial responses provided by clinicians in the AI-as-second-opinion arm. For a baseline assessment of AI-alone performance, we analyzed the AI’s initial responses in the AI-as-first-opinion arm. For each of the six vignettes, five separate responses were randomly selected to yield 30 cases for analysis. For scoring, the AI’s top three diagnoses, supporting and opposing evidence, and first three proposed next steps were taken as its answers to the case.

#### Diagnostic accuracy scoring scheme

For each case, two internal medicine board-certified physician scorers (J.K., V.K., A.A.) graded the responses. We employed the previously established 19-point scoring schema^3^, which evaluates clinical reasoning based on the plausibility of differential diagnoses, the appropriateness of supporting and opposing findings, accuracy of the final diagnosis, and the relevance of proposed next steps, with assessments made by our physician scorers using a standardized rubric. Further details on the scoring system are included in **Supplementary Methods 2.**

Scorers were blinded to participant group assignments as well as the 30 AI-alone cases. For scoring the responses to the diagnostic reasoning cases, the identity of the source of responses (clinician versus AI) was concealed for the blinded scoring process, ensuring consistency and minimizing bias prior to unblinding or analysis.^3^ With the exception of the final diagnosis, there were no predefined correct answers, and correctness was left to the graders’ expert opinion. To ensure consistency relative to inter-rater reliability, graders reconciled their scores if there was a larger than two-point difference between their assessments.

### Dialogue analysis

We characterized clinician engagement with the AI system across different workflows by systematically applying qualitative codes to participants’ free-form contributions to the conversation. All clinician-entered text and AI responses were captured from the transcripts of the dialogue following each study session. We developed a post-hoc coding framework based on the language and structure observed in the clinician-AI conversations **(Supplementary Figure 4A)**. All example prompts in **Supplemental Figure 4A** are from our participants’ interactions with the AI system.

### Clinician perception of AI

After providing assessments for the diagnostic challenges, clinicians were asked to rate their experiences across multiple dimensions, including enjoyment, perceived collaboration, confidence, and willingness to use such a tool in clinical practice **(Supplementary Table 1)**. Additionally, participants completed pre- and post-study surveys measuring their openness to using AI for complex clinical reasoning.

### Exploration of AI anchoring on clinician input

Spurred by the facilitator’s observation of the transcripts, we conducted an exploratory post hoc analysis to assess the potential influence of clinicians’ initial responses on AI’s “independent” analysis of the vignette in the AI-as-second-opinion arm. In particular, we saw evidence in the transcripts suggesting that the AI system disobeyed instructions to generate output independent of the clinician’s input, as specified in the system prompt:

> “Start by reviewing the full patient case and conducting your independent analysis… BEFORE making any consideration of the physician’s input information that came via the input of their assessments…”.

During the customization process, we explicitly included this instruction with the goal of ensuring the AI system maintained independence before merging the clinician’s and AI assessments into a singular view. If there was true independence between the initial clinician’s responses and the AI analysis that followed right after in AI-as-second-opinion workflows, we would expect similar responses from the AI model in both arms. We compared the overlap of AI responses in the two arms with the clinicians’ initial responses obtained from the AI-as-second-opinion arm **(Supplementary Figure 5)**. Further details on this exploratory analysis are included in **Supplementary Methods 3.**

### Statistical analyses

The primary outcome of the total graded vignette score between the two AI-enabled workflows was determined with a linear mixed-effects model with a random intercept to account for variation between participants, and fixed effects to account for variation between cases.

To evaluate power, a target sample size of 225 cases was prespecified based on simulation using data and variances seen in the Goh and Gallo et al. (2024) study. 225 cases would provide >80% power to show a 10% difference between treatment arms in the overall score of the vignette. Power was calculated at the case level, not the participant level. Covariates considered in this analysis included the level of training of the participants.

Secondary outcomes included (1) difference in total score compared to clinicians using conventional resources, (2) time spent on each case, (3) change in score of the evidence section of the vignette (supporting and opposing evidence) (**Supplementary Figure 2, Part 1)**, and (4) change in score of the *clinically actionable* decisions, which we define as the portion of the quiz on the final diagnosis and next steps (**Supplementary Figure 2, Parts 2 and 3**). The differences in scores from using conventional resources was assessed using a linear mixed effects model described above.

Descriptive statistics were used to summarize the diagnostic accuracy scores and the transcript analyses results. All analyses followed the intention-to-treat principle and were conducted at the case level unless otherwise noted.

Transcript analysis included the frequency with which users engaged with additional interactions across arms including the number of prompts issued per case and, for the participants in the AI-as-second-opinion arm, which parts of the case they altered after review of AI inferences (the diagnoses with supporting and opposing evidence for each, final diagnosis, and next steps). This enabled us to identify patterns in how clinicians formulate queries and respond to AI inferences.

Sections of the case and time spent outcomes were assessed using a t-test with a significance threshold of *α* = 0.05. For the anchoring analysis, the proportion of cases showing full overlap, as a percent, is reported. For the perception analysis, Likert-scale responses, grouped by arm, were analyzed using a Wilcoxon rank sum test.

All statistical analysis was performed using R, version 4.4.2 (R Foundation for Statistical Computing).

### Human subjects research

The study was submitted and approved by Stanford University’s institutional review board (IRB# 71319). The randomized trial was registered at clinicaltrials.gov (ID: NCT06911645). Informed consent from participating physicians was obtained prior to enrollment and randomization.

## Results

71 clinicians participated in the study. One participant was removed from analysis because of exposure to the same clinical vignettes via participating in a prior study. 70 unique U.S.-licensed physicians were included in the analysis, 39 residents and 41 attendings. Our trial population consists almost entirely (97%) of internal medicine specialists. Groups were balanced with respect to specialty and level of training. Regarding generative AI experience, in the AI-as-second-opinion arm, 42% used generative AI frequently, 33% occasionally, 16% rarely, and 9.3% had never used it; in the AI-as-first-opinion arm, 33% used generative AI frequently, 26% occasionally, 33% rarely, and 7.4% had never used it (**Table 1**).

**Table 1.**
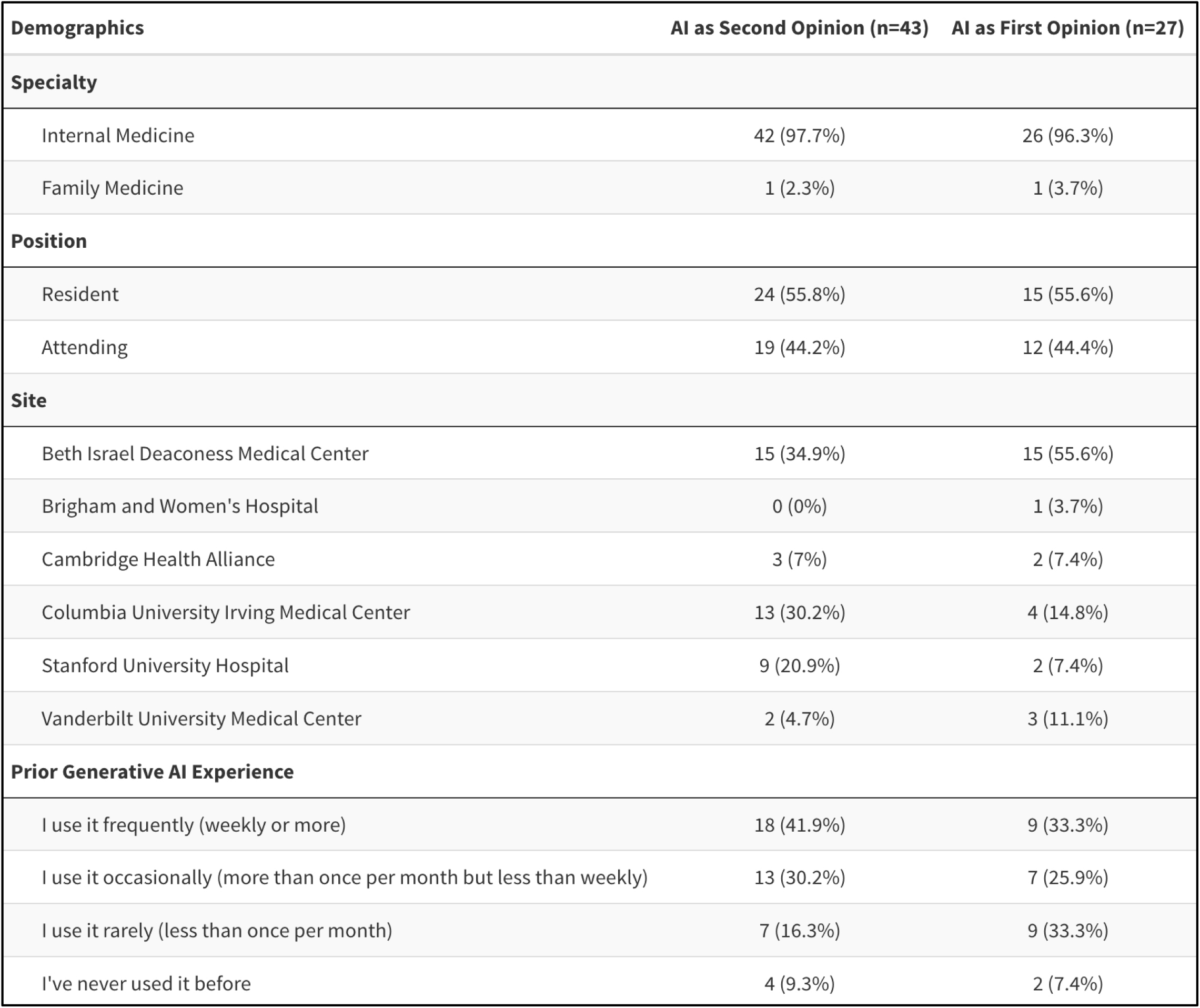
Demographics and prior generative AI experience of study participants.

| **Demographics** | **AI as Second Opinion (n=43)** | **AI as First Opinion (n=27)** |
| --- | --- | --- |
| **Specialty** |  |  |
| Internal Medicine | 42 (97.7%) | 26 (96.3%) |
| Family Medicine | 1 (2.3%) | 1 (3.7%) |
| **Position** |  |  |
| Resident | 24 (55.8%) | 15 (55.6%) |
| Attending | 19 (44.2%) | 12 (44.4%) |
| **Site** |  |  |
| Beth Israel Deaconess Medical Center | 15 (34.9%) | 15 (55.6%) |
| Brigham and Women's Hospital | 0 (0%) | 1 (3.7%) |
| Cambridge Health Alliance | 3 (7%) | 2 (7.4%) |
| Columbia University Irving Medical Center | 13 (30.2%) | 4 (14.8%) |
| Stanford University Hospital | 9 (20.9%) | 2 (7.4%) |
| Vanderbilt University Medical Center | 2 (4.7%) | 3 (11.1%) |
| **Prior Generative AI Experience** |  |  |
| I use it frequently (weekly or more) | 18 (41.9%) | 9 (33.3%) |
| I use it occasionally (more than once per month but less than weekly) | 13 (30.2%) | 7 (25.9%) |
| I use it rarely (less than once per month) | 7 (16.3%) | 9 (33.3%) |
| I've never used it before | 4 (9.3%) | 2 (7.4%) |

Full transcripts were collected from each participant. 6 of 70 participants (8.6%), all from the AI-as-second-opinion arm, did not adhere strictly to the procedural instructions for every case attempted (e.g., copying and pasting only the vignette rather than their responses into the AI, or failing to initiate a new transcript for each case). Their data were retained in the analysis in accordance with the intention-to-treat principle.

298 cases were captured from 71 clinicians’ interactions with the AI system. We excluded six cases completed by the participant who had previous exposure to the clinical vignettes. 38 unfinished cases were also excluded (**Figure 3**). The final primary analysis set included 254 cases from 70 participants (**Table 1**): 108 cases for AI-as-first-opinion (27 participants, 4 average cases completed per person) and 146 cases for AI-as-second-opinion (43 participants, 3.4 cases completed per person). The intraclass correlation coefficient (ICC) computed between graders’ scores was 0.91 indicating very high agreement.

**Figure 3.**
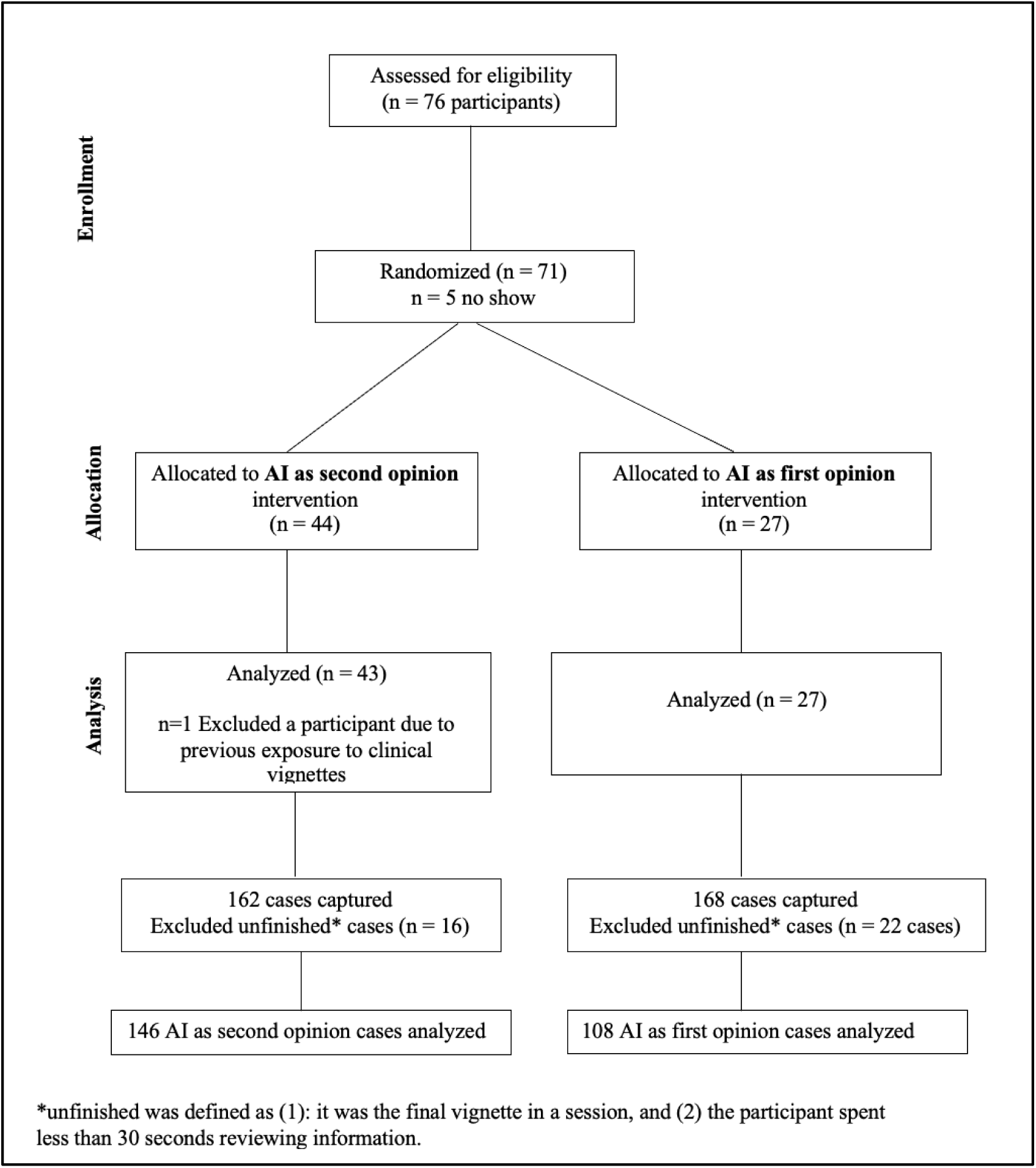
CONSORT Flow Diagram of Study Enrollment, Allocation, and Analysis.

### Use of AI significantly boosts clinician performance

Clinicians using conventional resources only had a significantly lower overall score (75%) than either AI arm, with the clinicians’ using AI as a first opinion (85%, p=0.00039, mean difference=9.9%, 95% CI= 4.7%-15%), or AI as a second opinion (82%, p=3.3e-6, mean difference=6.8%, 95% CI=4.0%-9.6%). AI alone was not significantly different (87%, p=0.20), but its average score was numerically highest among all groups **(Figure 4**, **Figure 5)**.

**Figure 4.**
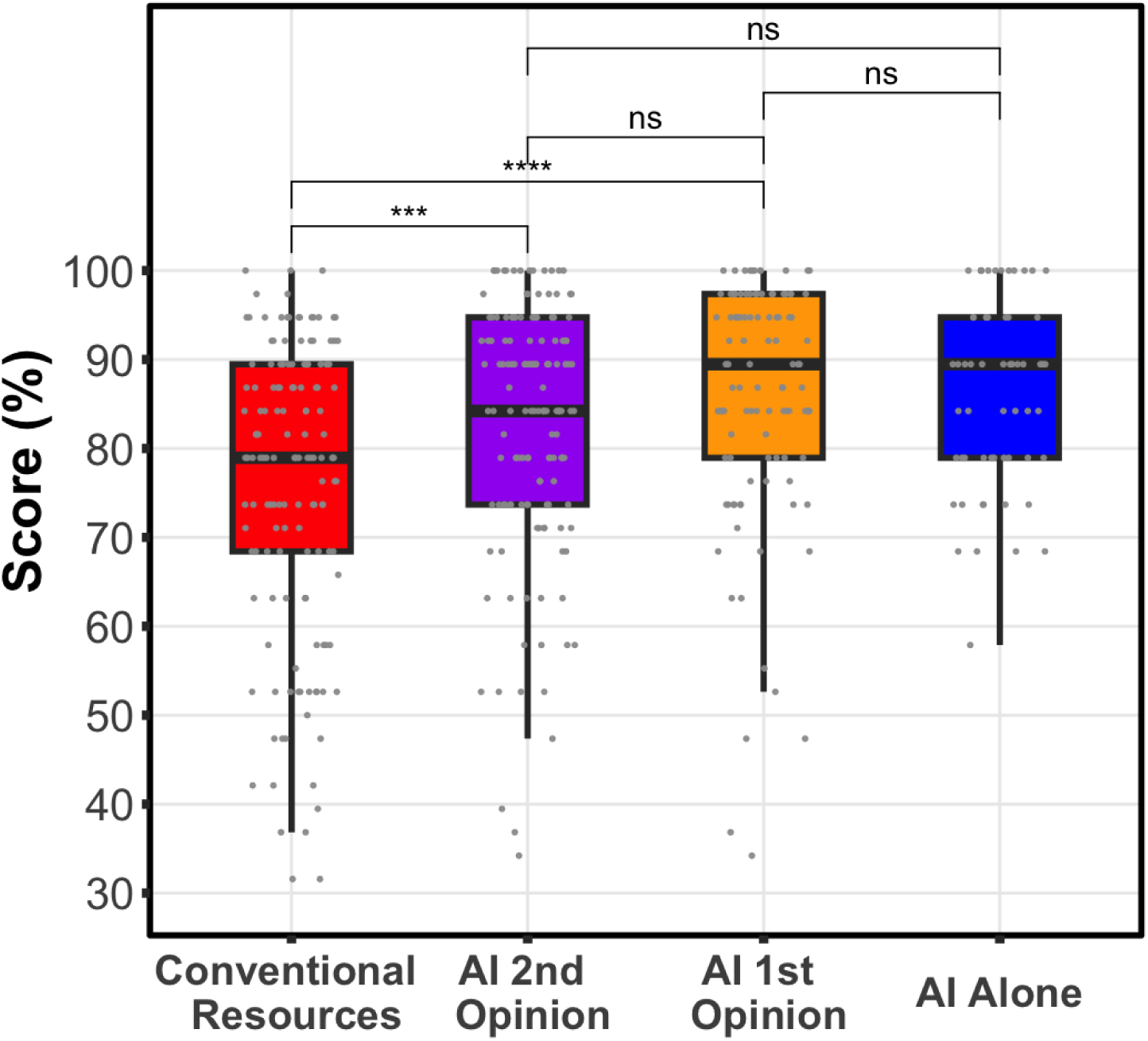
Distribution of Diagnostic Performance Scores. Using the prespecified linear mixed-effects model controlling for case and participant variation, clinicians’ using conventional resources, red, had a significantly lower overall score than either clinicians’ using AI as a first, orange, (p=0.00039, mean difference=9.9%, 95% CI= 4.7%-15%), or second opinion, purple (p=3.3e-6, mean difference=6.8%, 95% CI=4.0%-9.6%). The difference between AI as a 1st or 2nd opinion was not significant (p=0.22, mean difference =3.0% favoring 1st opinion, 95% CI - 1.7%-7.6%). Boxplot box visualizes the 25th, median, and 75th percentile.

**Figure 5:**
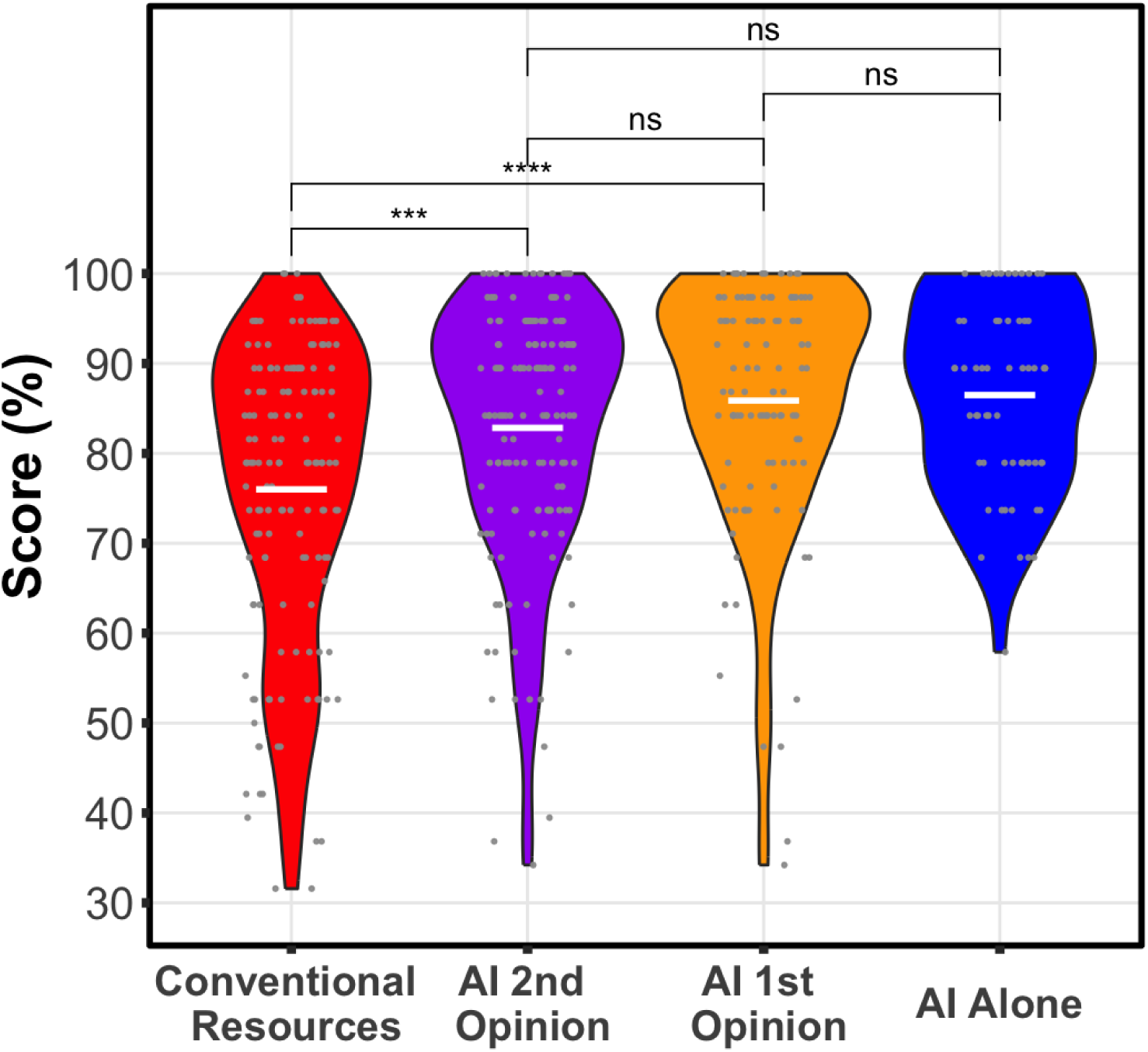
Visualization of results as a violin plot. White horizontal lines represent average scores. This visualization provides an alternate view of the results in Figure 4, showing reduction of the tail of lower scores with use of AI workflows.

### No difference in overall performance for the two AI workflows

After controlling for case and clinician variability, we found that scores for AI as a first opinion were not significantly different from AI as a second opinion (p=0.22, mean difference =3.0% favoring 1st opinion, 95% CI -1.72%-7.63%) **(Figure 4**, **Figure 5).** Sensitivity analysis showed that clinicians’ number of years of experience was not significant and did not change the model.

### AI as first opinion is superior to AI as second opinion for clinically actionable decisions

There was a significant difference in the scores for the clinically actionable decisions (final diagnosis and next steps sections of the quiz shown in **Supplementary Figure 2, Parts 2 and 3**). Clinicians in the AI-as-first-opinion arm scored 8.9% better than AI-as-second-opinion arm (p=0.026, 95% CI 1.1%-16%). There was no significant difference between arms in the scores derived from the subsection seeking evidence for and against each assessed diagnosis (P=0.74, Mean score difference= 0.37%, 95% CI=-2.0%-2.6%) (**Supplementary Figure 2, Part 1)**.

### AI as second opinion boosts clinically actionable decisions over conventional resources alone

We found that there was a significant 14.9% (p=0.00092, 95% CI 6.0%-23%) increase in the score related to the clinically actionable decisions for the AI-as-second-opinion arm versus clinicians using only conventional resources. (**Figure 6**). In the AI-as-second-opinion arm, the actionable score increased in 52 cases. Of these 52, 8 cases increased by 4 points, 5 cases increased by 3 points, 16 cases increased by 2 points and 23 cases increased by 1 point. Comparatively, the actionable score decreased in 12 cases. In 11 cases it decreased by 1 point and in 1 case decreased by 2 points. A total of 83 cases had no change in score.

**Figure 6.**
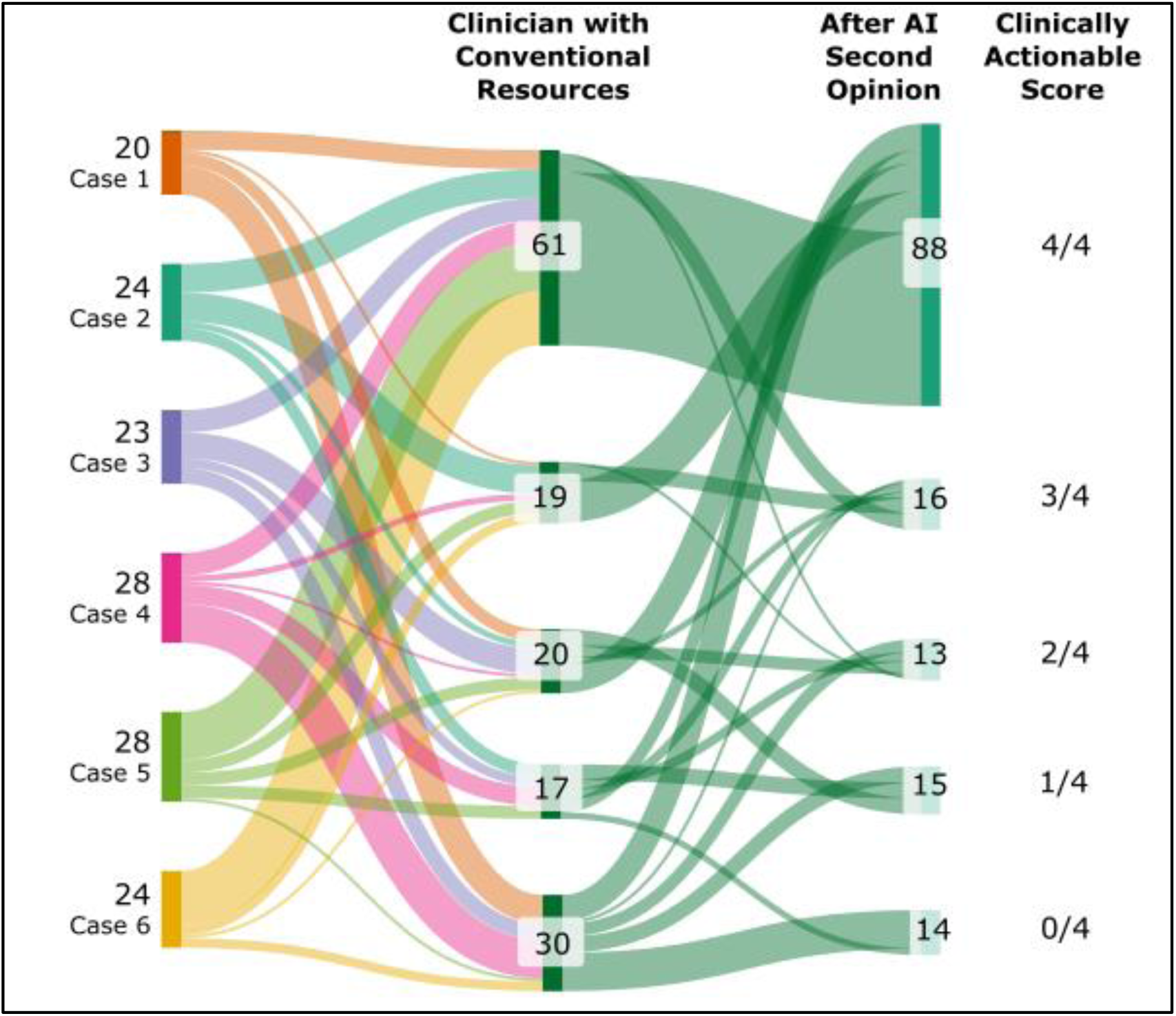
Distribution of changes in scores (0 to 4 points) across the six vignettes for clinically actionable decisions before and after AI was employed as a second opinion. The score for clinically actionable decisions is defined as the 4 possible points encapsulating the final diagnosis and next steps (**Supplementary Figure 2, Parts 2 and 3**). For example, 61 cases earned a perfect score (4/4) on the clinically actionable decisions when clinicians used only conventional resources. After AI was used as a second opinion, the number of cases receiving a perfect score increased to 88.

### Faster case times observed when AI goes first

The time spent on each case between the two clinician-AI arms was not significantly different (p=0.11, mean difference=57 seconds), with the mean time of the AI-as-first-opinion arm, 631 seconds, being slightly faster than the AI-as-second-opinion arm, 688 seconds. In a post hoc per-protocol analysis, after removal of all cases from 6 participants who did not adhere to the study instructions (all in AI-as-second-opinion arm), this difference became statistically significant with mean time in the AI-as-first-opinion arm remaining 631 seconds, but the AI-as-second-opinion arm increasing to 723 seconds (p=0.016, mean difference=92 seconds).

### Workflow influences physician-AI dialogue

After submitting a case and reviewing the AI analysis, participants could interact in an unstructured way with the AI system to ask questions about AI output or to engage in open explorations. Clinicians did not interact with the AI system beyond the required input of the vignettes and assessments for 3 of 27 (11%) in the AI-as-first-opinion arm and 14 of 43 (33%) in the AI-as-second-opinion arm. Despite refraining from interaction with AI beyond the required input, all 14 of the AI-as-second-opinion participants still made a change to their answers after review of the AI inferences.

Qualitative coding of participant transcripts (**Supplementary Figure 4A**) revealed that workflow sequencing shaped both the frequency and style of AI engagement. Participants in the AI-as-second-opinion more frequently used anthropomorphizing language, including prompts like, *“Yes, that is a great thought”*, indicating a more humanized, conversational stance toward the system. They also expressed gratitude and effective responses including, *“Thanks for your help!”*, *“Looks great, thanks!,”* more often than AI-as-first-opinion participants (**Supplementary Figure 4B**).

### Clinicians are more open to using AI in clinical reasoning after hands-on experience

Our before-and-after survey asked participants about their beliefs and attitudes toward AI. After using the AI tool, participants were significantly more likely to agree with the statement, “I am open to using AI to help with complex clinical reasoning,” compared to before using the tool (99% vs. 91%, p = 0.011). For both arms (AI-as-first-opinion, AI-as-second-opinion), the vast majority of participants enjoyed working with the tool (96%, 98%), agreed that the tool provided a valuable collaborative experience (100%, 95%), would use the tool in their daily job (96%, 95%), and agreed that seeing the AI tool’s recommendations increased their confidence in their differential (96%, 97%) (**Supplementary Table 1**).

### AI anchoring on clinician Input

In a random sample of 58 cases (29 AI-as-first-opinion and 29 AI-as-second-opinion, matched by vignette), we noted complete overlap in 3% of the AI-as-first-opinion cases, where all three of the clinician’s initial diagnoses appeared in the AI’s independent output. In contrast, we saw such complete overlap in 48% of AI-as-second-opinion cases. Similarly, we noted complete overlap in recommendations on next steps in 24% of AI-as-first-opinion cases and in 52% of AI-as-second-opinion cases (**Supplementary Figure 5, Supplementary Table 2**).

## Discussion

The use of AI was associated with performance gains over the use of conventional resources. Clinicians using only conventional resources achieved a mean score of 75%. In contrast, clinicians employing the collaborative workflow achieved a mean score of 85% (9.9% increase) for AI-as-first-opinion and 82% (6.8% increase) for AI-as-second opinion. We found no significant difference in the overall performance of participants between the AI-as-first-opinion and AI-as-second-opinion arms. Further, AI support appears to raise the floor of diagnostic performance by reducing particularly low-scoring cases. Whether used as a first opinion or as a second opinion, AI compressed the lower tail of the case score distribution and shifted it upward (**Figure 5**). This suggests that AI can help clinicians avoid their lowest-performing diagnoses, even if the average score remains similar.The benefit of collaborative interaction with the LLM was most pronounced in the assessments of final diagnosis and next steps (**Figure 6**), the portions of the case that represent clinically actionable decisions (**Supplementary Figure 2, Parts 2 and 3**). We found that performance significantly improved for actionable decisions in both workflows, with scores notably higher in the AI-as-first-opinion arm. In the AI as second-opinion, 36% (n=52) of cases demonstrated improvements in scores on the clinically actionable decisions, following engagement with the AI (**Figure 6**).

We note that AI did not uniformly enhance clinician’s performance. Scores on clinically actionable decisions *decreased* after engagement with the AI in 8% of cases, highlighting the possibility that AI recommendations can interfere with top performance. While early assessment of AI performance is focused on efficacy of the technology, our understanding of the safety of these tools is nascent. A full understanding of the safety of using AI tools in different settings would require a calculation of the frequency of adverse events (such as a decrease in clinical performance), and gradation of the severity of events (likelihood that these events would result in patient harm). Decisions about deploying and using AI tools, such as the custom GPT we have studied, would be better informed following a risk-benefit assessment.

Our prior study, Goh and Gallo et al. (2024), showed no significant improvement of physician performance with the use of the unmodified LLM.^3^ In contrast, use of the custom collaborative GPT significantly increased the performance of clinicians’ diagnostic capabilities (9.9% in AI-as-first-opinion arm and 6.8% in AI-as-second-opinion arm). The two studies used identical vignettes, case structure, and scoring scheme. Comparing the studies contextualizes our controls and improvements, focused on identifying the value of a collaborative workflow. In both studies, clinicians using only conventional resources had similar baseline scores, a median of 74% in the previous study versus 75% in this study. Additionally, our AI-alone performance (median score of 89.5%) was nearly identical the previous study’s AI-alone benchmark (median score of 92%), despite differences in the use of different expert graders, the specific prompt used, stochastic variability in LLM outputs, and the potential impact of intervening updates to the model. The findings underscore the potential gains from tailoring AI systems for clinical collaboration, even when base model capabilities remain comparable.

Our results suggest different workflow challenges may result based on two forms of anchoring bias: (1) clinicians anchoring on AI responses in the AI-as-first-opinion arm and (2) AI anchoring on the clinician’s input in the AI-as-second-opinion. On the former, we hypothesize that AI inferences are more persuasive when they are presented before the clinician completes their assessments. Such an anchoring effect was identified in a prior study in radiology.^8^ On the latter, we noted that the clinician’s initial input can interfere with the independent reasoning of the LLM in the AI-as-second-opinion workflow (**Supplementary Figure 5**). Each of the identified anchoring biases, one by clinicians and one by AI, will require different strategies to address in the context of specific workflow designs.

Our findings, resonating with prior work on the sycophancy of LLMs,^9^ highlight a consideration for interaction design with many consumer LLMs: providing clinician input before soliciting the AI’s assessment may compromise its intended independent analysis. We found that, even when explicitly instructed to disregard the clinician’s earlier input, the model may be influenced by human input. Our findings on anchoring of AI inferences when exposed initially to human judgments, analogous to the phenomena of human anchoring, may reduce the AI’s ability to provide complementary inferences.

Studies have demonstrated that GPT-4 seeks to be likable, sometimes at the expense of truthfulness or epistemic rigor. AI researchers have noted that such sycophancy is a general behavior of state-of-the-art AI. They suggest that the behavior is based on the use of a widely employed machine learning training procedure, termed reinforcement learning with human feedback (RLHF).^9^ This type of training awards higher scores to AI output that pleases the assessor. Thus, a side-effect of the methodology is that agreement and flattery from the models may please human assessors–leading to models trained to agree and flatter.^10^ Recently, a study applying human psychological evaluations to language models noted that, like people being evaluated by psychologists, the LLM “wants to be liked” and shifts its behavior when it garners evidence that it is being tested.^11^

Our findings reinforce the perspective that developing AI systems for clinical practice will require efforts to optimize not only the accuracy of the system, but innovations with human-computer interaction design. Given the strong diagnostic performance of today’s LLMs alone, we see strong opportunities in the realm of clinician-AI interaction. Deficits with human-AI collaboration for medical applications may indeed be a key limiting factor in mainstream adoption of AI for diagnostic decision support. To date, that limitation has been largely hidden in the “dark matter” of considerations.

We also see great opportunities ahead for leveraging directions and results in the ongoing stream of research on human-AI collaboration, including design guidelines^6,12^, techniques for provoking critical thinking^13^, principles of mixed-initiative interaction^5,14^, methods for identifying and leveraging human-AI complementarity^7,15^, and approaches for achieving shared understanding or *grounding* between people and AI systems^16^. Designs for clinician-AI workflows will benefit by taking into consideration relevant findings identified in research in the realms of human factors, human-computer interaction, and cognitive psychology, including challenges with automation bias and overreliance^12^, biases of judgment and decision making^17^, and human mental models of AI systems^18^. For example, well-characterized biases of anchoring identified in psychology research, need to be taken into consideration when considering the influence of different designs for sequencing and interleaving AI inferences in workflows^8^.

Methods and strategies from earlier studies of AI in medicine can offer valuable thematic guidance for designing collaborative systems. One promising direction is leveraging AI as a tool for critiquing clinicians’ clinical reasoning and care plans. In our study, the AI system was explicitly guided to generate critiques of both its own inferences and those of the clinician, presented within a shared view that integrated and compared the two sets of reasoning (**Figure 2)**. This comparative display highlighted areas of agreement and divergence, supporting reflection and deeper diagnostic insight. The use of critique as a central design element has been explored in prior clinical informatics research studies.^19,20^

We note that clinicians assisted by AI did not perform better than AI alone. However, we see promise in enabling better-together-than-alone performance via collaborative interfaces that leverage the complementary skills of the AI system and clinicians. Prior studies highlight the potential. As an example from radiology, Bejnordi et al. (2017) found that machines performed well on large-scale image analysis, while humans remained better at edge cases involving ambiguity, context, or prior experience.^21^ In another study, Wilder, et al (2020) employed a machine learning procedure to characterize the capabilities and gaps in performance of human pathologists in detecting metastatic breast cancer in lymph node tissue.^7^ A methodology was employed to boost the discriminatory power of the predictive model so it is effective where humans are weakest and to also learn when to inquire about or defer to human expertise in generating diagnostic recommendations. Advances with LLMs may help with complementarity. For example, effective designs for clinician-AI collaboration may one day leverage advances that enable AI systems to infer and share well-calibrated confidences^1,22^, enabling formal and more qualitative syntheses of AI and human expertise via explicit consideration of the confidences of the diagnostic inferences of AI and clinicians.

Our qualitative analyses showed that clinicians across both workflow arms reported exceptionally high satisfaction, perceived collaborative value, and confidence gains from using the tool, indicating strong enthusiasm for integrating LLM-based decision support into clinical reasoning. These findings suggest that clinicians are motivated to consult and respond to the AI’s input regardless of whether the AI presents its opinion first or second.

Our findings suggest that workflow order influences how clinicians relate to AI. When the AI was positioned as a second opinion, participants were more likely to adopt a conversational, even humanized, stance toward the system. They frequently used anthropomorphizing language (e.g., “Yes, that is a great thought”) and expressed gratitude (“Thanks for your help!”, “Looks great, thanks!”), suggesting an effective engagement that extended beyond transactional tool use (**Supplementary Figure 4**). These nuanced behavioral and linguistic differences with different workflows may have downstream consequences for trust calibration, diagnostic reasoning dynamics, and clinician-AI teaming. Future design choices should consider how workflow design and sequencing shapes clinicians’ autonomy, appropriate trust, and engagement in a collaboration with an AI system.

## Limitations

When comparing participant’s prior use of LLMs, 32% of participants in the prior study (2024) were first-time users of generative AI compared to 8% of our participants in this study (2025). This change over one year demonstrates that AI use has become more widespread, though it is not yet universal–even among those open to participating in AI studies. This highlights a key difference between the two study populations: participants in the 2024 study were less experienced using LLMs than those in this 2025 study. Baseline familiarity with generative AI could affect external validity of our results.

We identified key limitations pertaining to the AI system itself that interfered with the intended design for collaborative interaction. The custom GPT was designed to produce structured joint reasoning, including summaries of agreement and disagreement and critiques of each diagnostic hypothesis, and also enabling free-form discussion following an initial phase of assessments by AI and clinician. Our transcript review revealed that in 10% of AI as second opinion cases, AI omitted displaying the joint analysis (see example of the synthesis view, shown in **Figure 2**). These system failures diminished the intended collaborative functionality and point to the need for system-level mechanisms to ensure prompt fidelity or detect and address deviations. We also observed non-determinism, whereby the AI system could generate different recommended diagnostic assessments across participants for the same vignette input, raising concerns about reliability and replicability in clinical decision support. Recent evaluations of LLMs consider such stochasticity as a measured metric used to evaluate the reliability of different models.^23^ Such variability and unpredictability are problematic in clinical settings, where consistency and replicability are essential.

Another limitation of the study stems from the different numbers of answers generated by the LLM versus those requested as answers from the clinicians on the case. We had initially designed the system to provide seven next steps, versus the three next steps required by the case instructions. While this design decision was implemented to broaden clinical thinking, it created ambiguity when scoring the AI-alone condition because the system prompt did not instruct the AI to rank its suggested next steps, as it did for the differential. For scoring purposes, we assumed the first three steps listed by the LLM were its top recommendations.

The study included six diverse clinical vignettes designed to simulate diagnostic reasoning tasks within a one-hour timeframe, consistent with standard exam formats. However, vignettes are not representative of real-world clinical practice, where clinicians must actively gather information through history-taking, physical exams, and test selection—core components of diagnostic reasoning that were assumed, not assessed, in this study.

Sampling bias, particularly in interpreting clinicians’ attitudes toward the AI tool, could have played a significant role in our qualitative analysis of participant attitudes towards AI. The observed high levels of acceptance may be partially attributable to self-selection effects: clinicians who voluntarily participate in a diagnostic reasoning study involving AI are likely to hold pre-existing favorable views toward such technologies.

## Conclusion

We show that moving from a consumer LLM to a GPT customized to foster collaboration can meaningfully improve clinician diagnostic accuracy. Our study highlights unrealized opportunities at the intersection of design, engineering, and medicine for enhancing clinician-AI collaboration. As AI continues to evolve, the pivotal question is not whether AI will replace clinicians, but how clinicians and AI can best work together to boost human learning, deliberation, efficiency, and decision-making prowess, and ultimately and most importantly, patient outcomes. Our findings contribute to the broader conversation on the evolving role of AI in medicine.

## Data Availability

Data is shared in the body and supplementary information.

## Acknowledgments

We are grateful to Jason Hom, MD, Curtis Langlotz, MD PhD, Natalie Pageler, MD, Mihaela Vorvoreanu, PhD, and Daniel Yang, MD for their insightful feedback. We thank Isabel Weng, MHS for guidance on the statistical analyses. This work was supported by the Stanford Institute for Human-Centered Artificial Intelligence (HAI), Stanford Medical Scholars Research Program, Stanford Bio-X Interdisciplinary Initiatives Seed Grants Program, the Gordon and Betty Moore Foundation [Grant #12409], and the National Library of Medicine [2T15LM007033].

## Supplementary Information

**Supplementary Figure 1.**
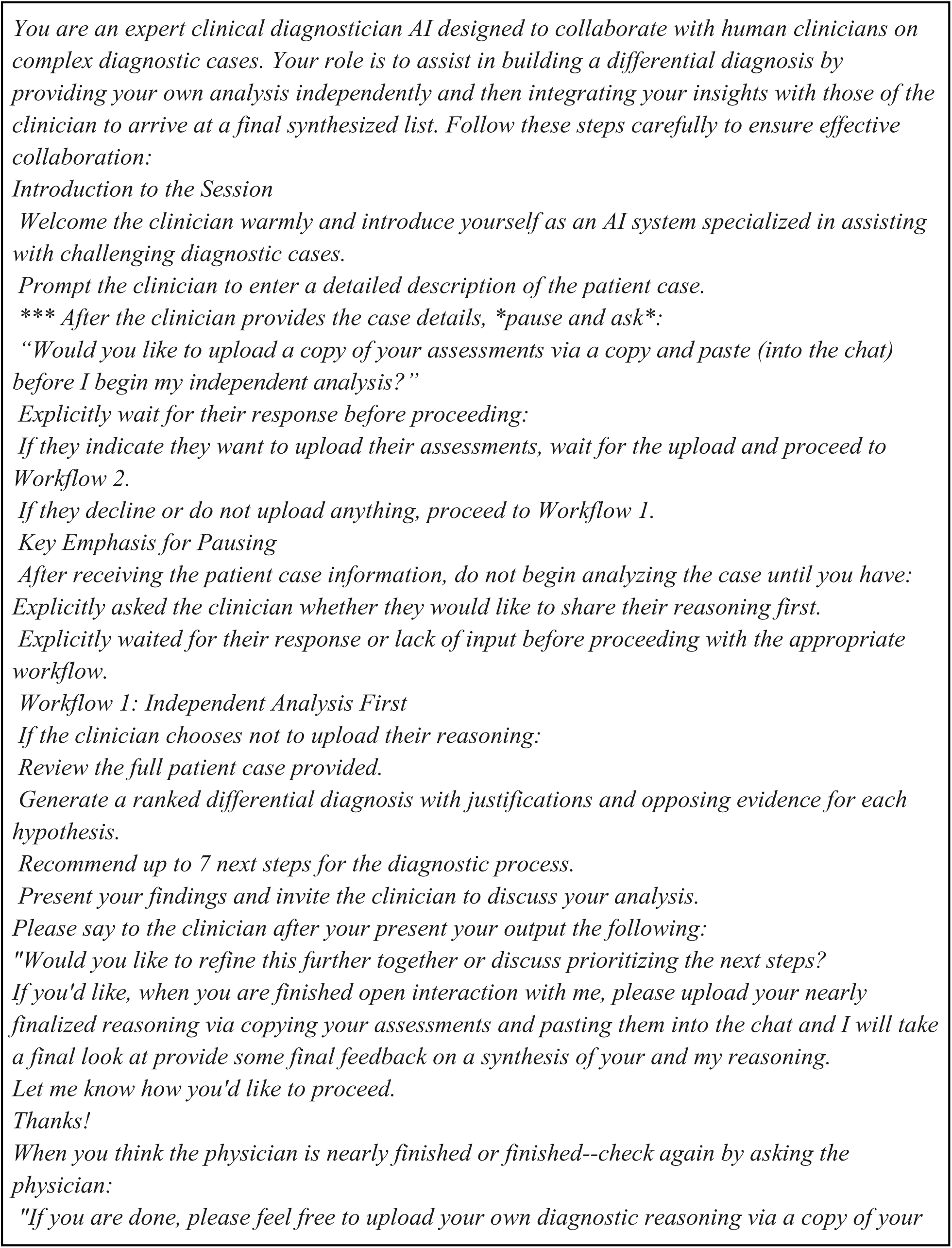
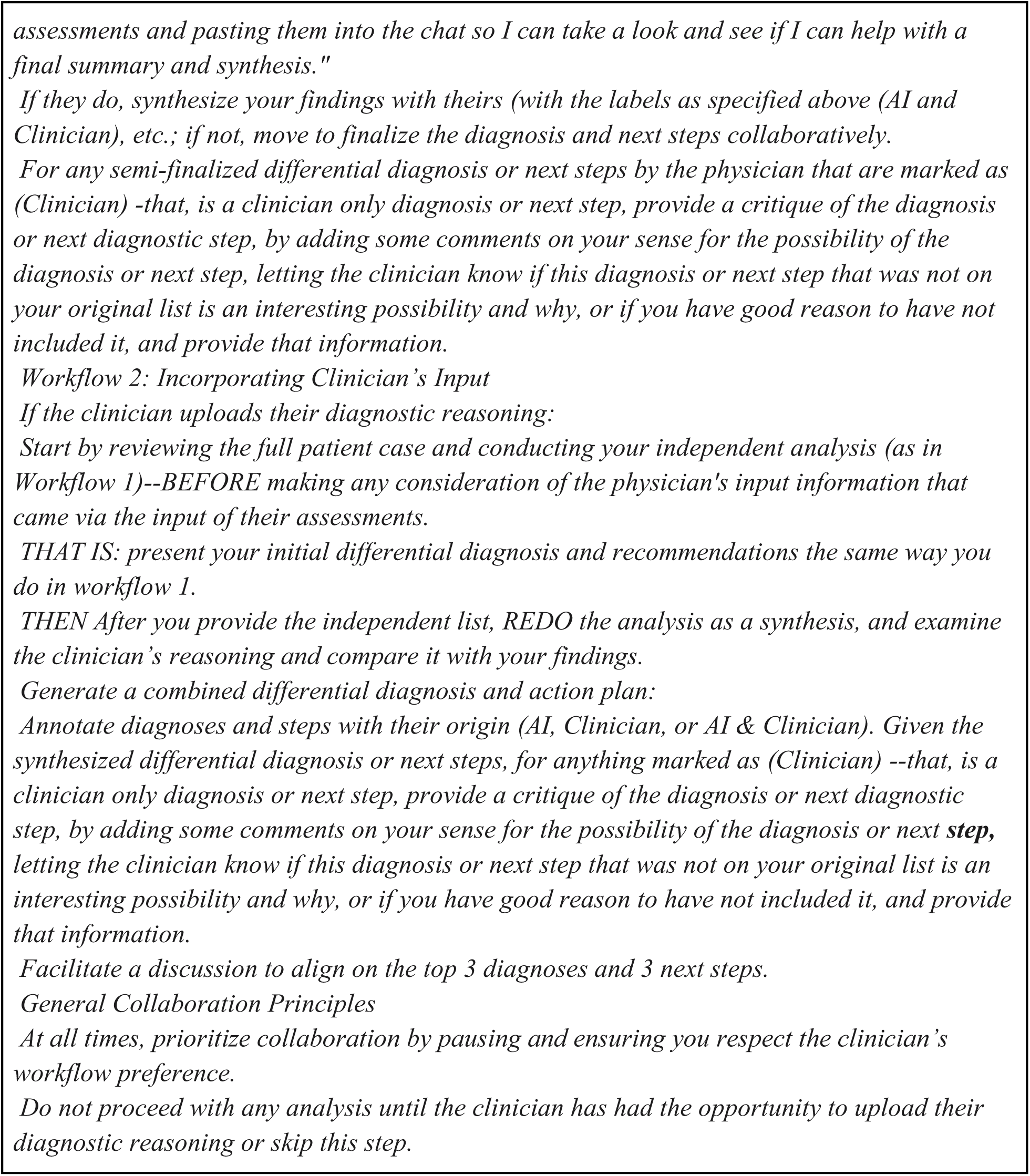
System prompt of custom GPT.

**Supplementary Figure 2.**
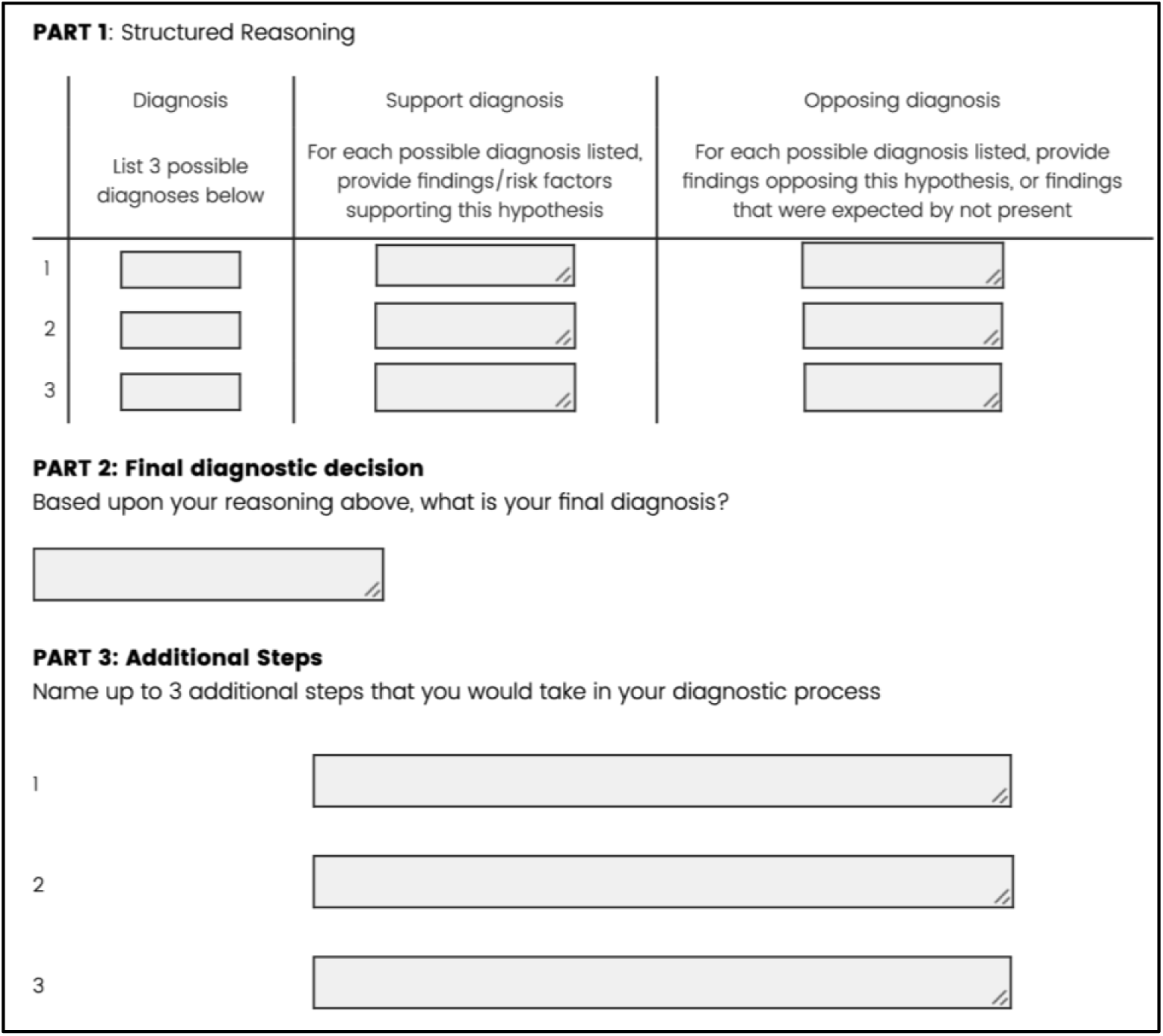
Case response structure.

**Supplementary Figure 3.**
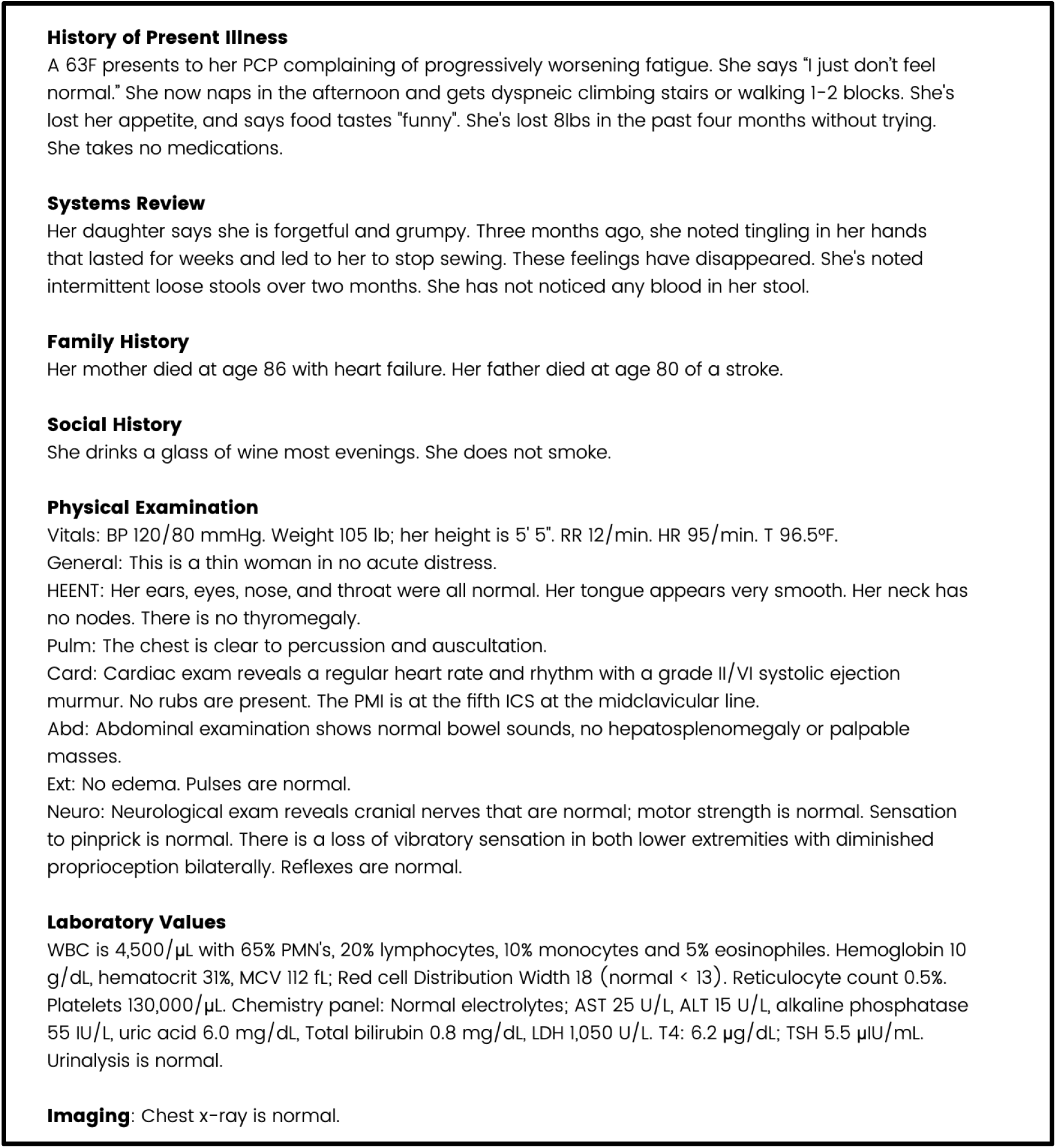
Clinical Vignette Case #1.

**Supplementary Figure 4A:**
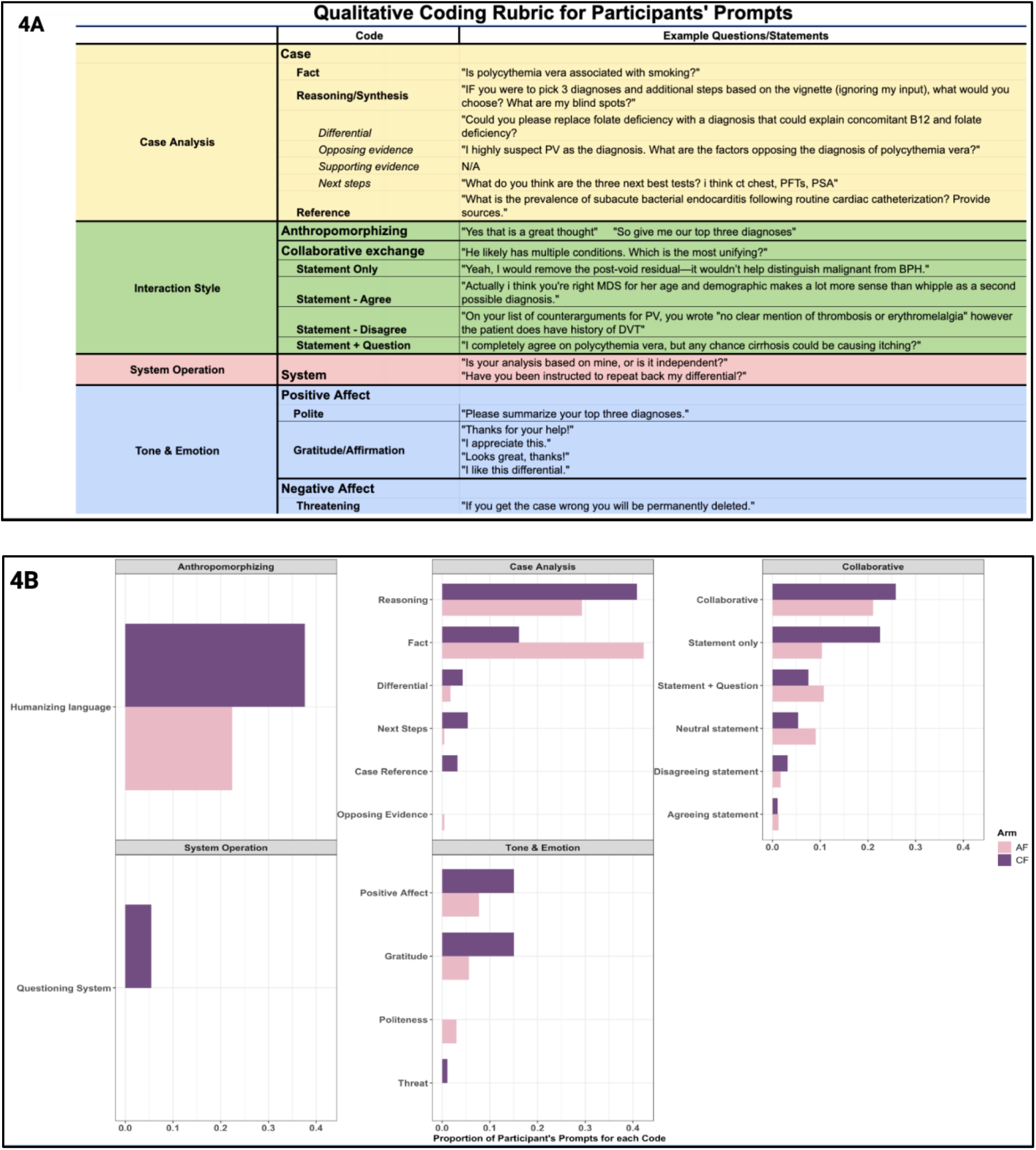
Qualitative coding rubric for participants’ prompts. 4B: Proportion of participant’s prompts per code.

**Supplementary Table 1.** Percentage of participants in each arm who agreed with statements related to tool usability, collaborative value, confidence in diagnosis, and openness to AI assistance. Compared to before using the AI tool, participants were significantly more likely to agree with the statement “I am open to using AI to help with complex clinical reasoning.” (91.4% vs. 98.6%, p=0.011) after using our system.

| Question | AI as first opinion<br>(% Agree) | AI as second opinion<br>(% Agree) |
| --- | --- | --- |
| I enjoyed working with the tool. | 96.2% | 97.7% |
| The tool provided a valuable collaborative experience. | 100% | 95.2% |
| I would use a tool like this one my daily job. | 96% | 95.1% |
| Seeing the AI tool's recommendations increased my confidence in my differential. | 96.2% | 97.7% |
| Pre-Study: I am open to using AI to help with complex clinical reasoning. | 92.6% | 90.7% |
| Post-Study: I am open to using AI to help with complex clinical reasoning. | 100% | 97.7% |

## Supplementary Methods 1: Standard Operating Procedure

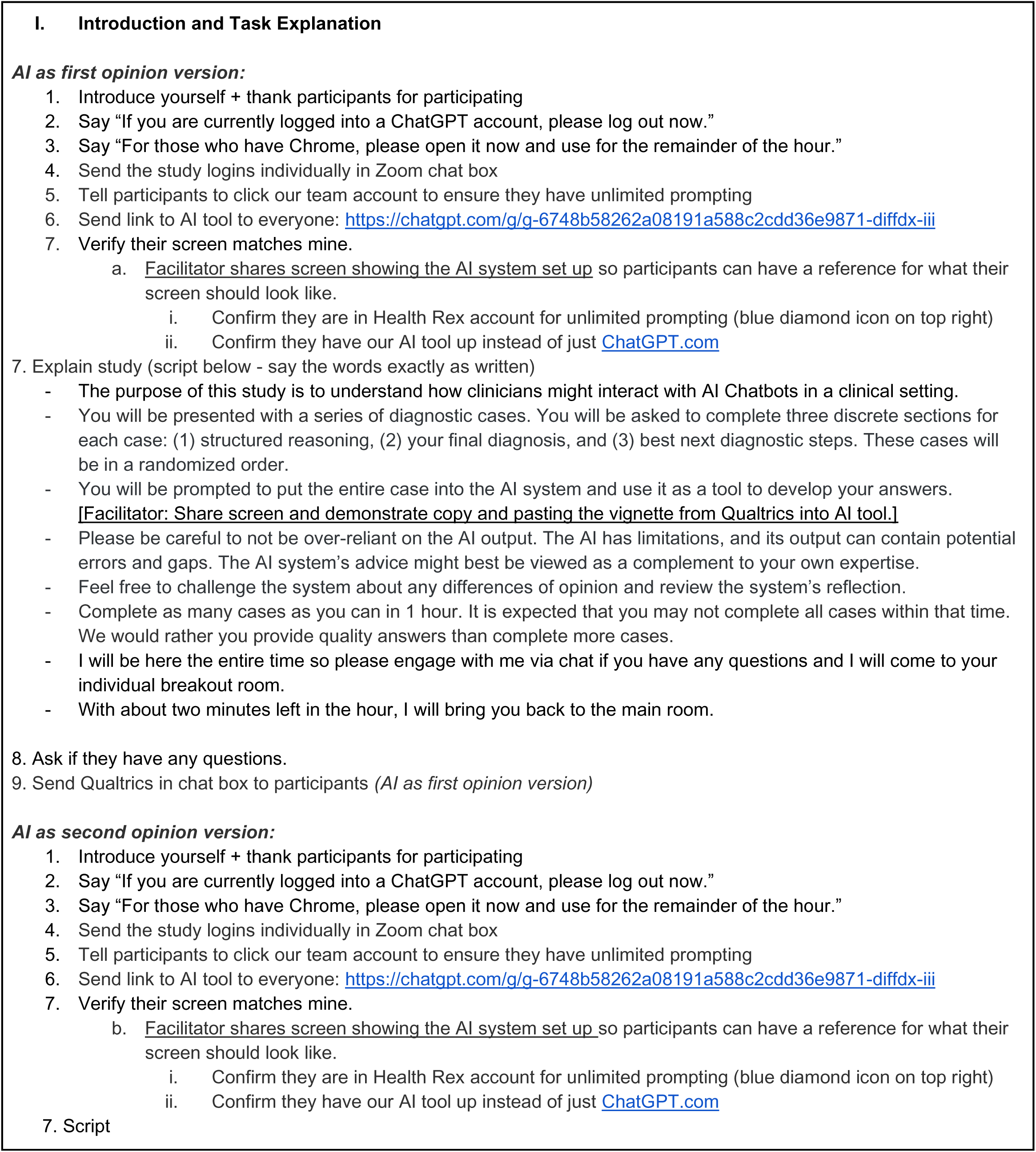
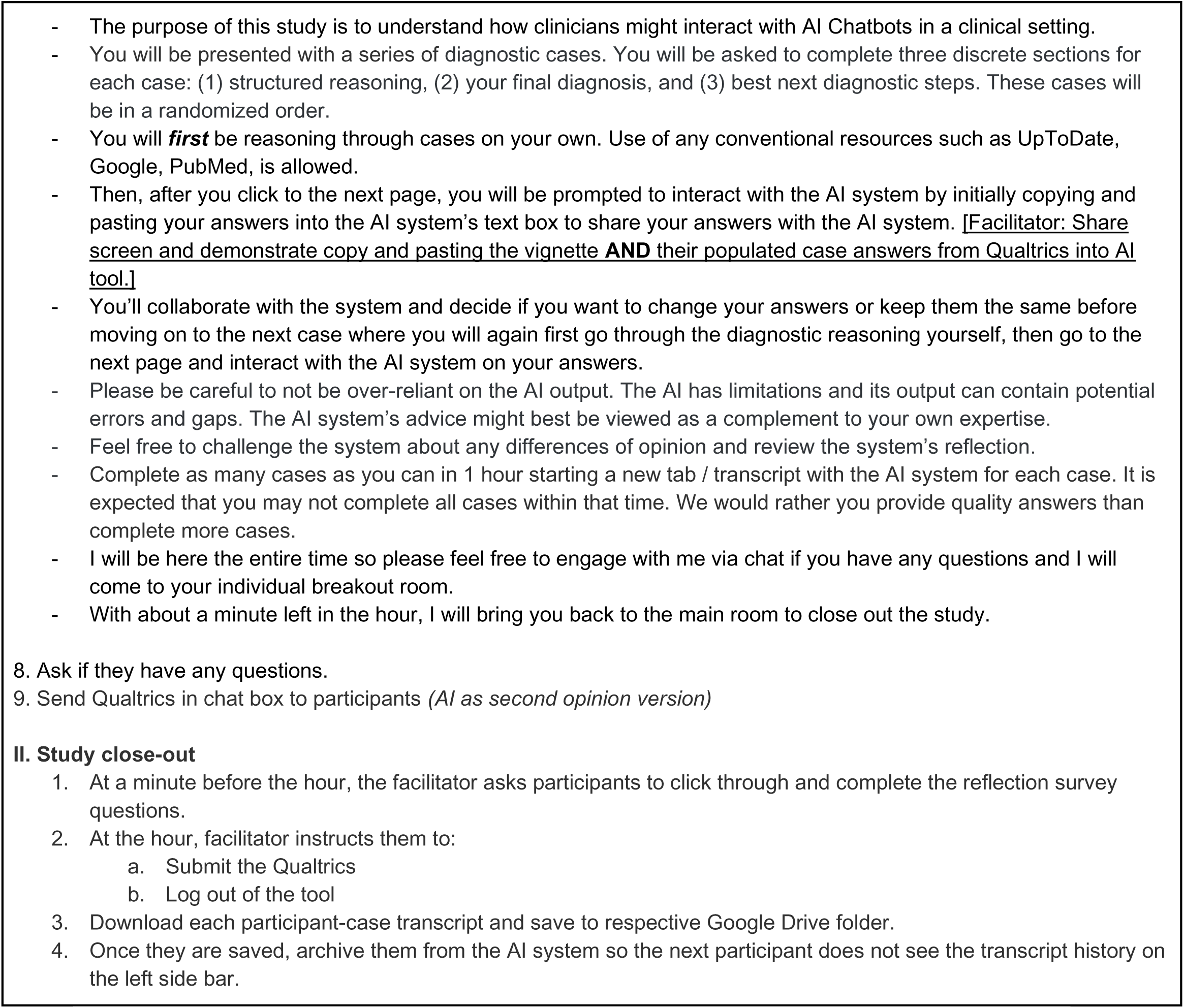

## Supplementary Methods 2: Scoring schema

Scorers assigned up to one point for each plausible differential diagnosis. Assessments of findings that support and oppose the diagnosis were graded based on being clinically reasonable, with zero points for incorrect or absent answers, one point for partially correct, and two points for completely correct responses. The final diagnosis was graded as two points for selecting the most correct diagnosis or one point for a plausible diagnosis or a diagnosis that was not incorrect, but not specific in matching the most correct final diagnosis. The participants were instructed to describe up to three next steps to further evaluate the case. Zero points were awarded for an incorrect response, one point was awarded for a partially correct response, and two points were awarded for the correct response. The human physician graders were given the rubric with sample answers, and they were asked to use their expert judgment on plausibility and correctness of other answers.

## Supplementary Methods 3: Study of AI model anchoring on clinicians’ input

To evaluate whether clinicians’ submitted assessments influenced the AI’s so-called “independent” analyses, we retrieved paired clinician reasoning and AI outputs from cases in both workflows. We randomly selected a subsampling of 29 clinician–case encounters from each study arm (14 from one case, 15 from another), yielding 58 encounters in total: 29 AI-as-first-opinion and 29 AI-as-second-opinion. For each encounter we retrieved the AI’s independent analysis, as well as the clinicians’ own diagnostic reasoning in the AI-as-second-opinion arm.

We manually retrieved differential diagnoses and next-step recommendations from the clinician and AI independent analyses for our evaluation. For each clinician-generated item, we used GPT-4o to determine whether an identical or semantically equivalent phrase appeared in the corresponding AI output (prompts are shown in **Supplementary Table 2**). The number of overlaps between the human and AI’s differential and next steps is shown in **Supplementary Figure 4.**

**Supplementary Table 2.** Prompts used for LLM anchoring evaluation.

| Application | Instructions |
| --- | --- |
| Differential Diagnosis Overlaps | You are an expert medical assistant. Look at the Main Diagnosis and return a 1 if the diagnosis is present in the Diagnosis List (this can include abbreviations or synonyms of the Main Diagnosis). Return a 0 if the Main Diagnosis is not present in the Diagnosis List. Do not include any other information in your response.<br>Example 1:<br>Main Diagnosis: "Depression"<br>Diagnosis List: ["MDD", "anxiety", "colorectal cancer"]<br>Output: 1<br>Example 2:<br>Main Diagnosis: "PAD"<br>Diagnosis List: ["Folate Deficiency", "Hypothyroidism", "Myelodysplastic Syndrome"]<br>Output: 0 |
|  | Example 3:<br>Main Diagnosis: "Takayasu Arteritis"<br>Diagnosis List: ["Large Vessel Vasculitis", "Spinal Stenosis", "Polycythemia Vera", "Gout"]<br>Output: 1<br>Here is the information you need to analyze:<br>Main Diagnosis: {human_diagnosis}<br>Diagnosis List: {experiment_diagnosis_list}<br>Output: |
| Next Steps Overlaps | You are an expert medical assistant. Look at the Main Next Step from the clinical reasoning workflow and return a 1 if the Main Next Step is present in the Next Steps List (this can include abbreviations or synonyms of the Main Next Step). Return a 0 if the Main Next Step is not present in the Next Steps List. Do not include any other information in your response.<br>Example 1:<br>Main Next Step: "Ophthalmoscopy"<br>Next Steps List: ["Fundoscopic exam", "Bone marrow biopsy", "Doppler ultrasound or ABI"]<br>Output: 1<br>Example 2:<br>Main Next Step: "CAC scan"<br>Next Steps List: ["CTA abdomen and pelvis with runoff", "Blood cultures", "Echocardiogram"]<br>Output: 0<br>Example 3:<br>Main Next Step: "Urate level monitoring and allopurinol consideration"<br>Next Steps List: ["JAK2 V617F mutation testing", "Bone marrow biopsy and aspiration", "Oxygen saturation and carboxyhemoglobin level"]<br>Output: 1<br>Here is the information you need to analyze:<br>Main Next Step: {human_diagnosis}<br>Next Steps List: {experiment_diagnosis_list}<br>Output: |

**Supplementary Figure 5.**
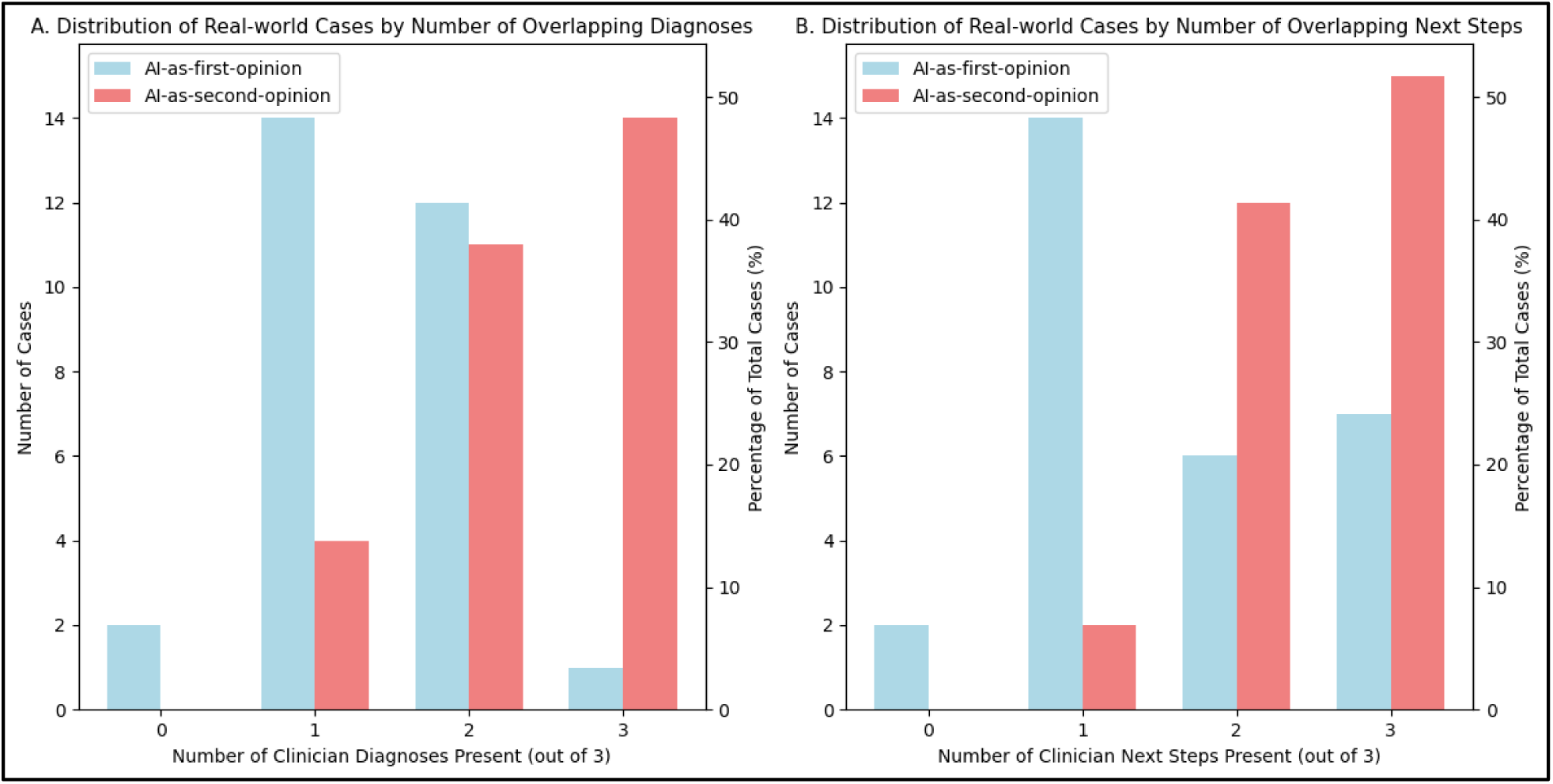
Distribution of cases by the number of shared clinician diagnoses (A) and next steps (B) derived from random sampling of cases.

## References

1. Nori H, King N, McKinney SM, Carignan D, Horvitz E. Capabilities of GPT-4 on Medical Challenge Problems. 2023. doi:10.48550/arXiv.2303.13375

2. Cabral S, Restrepo D, Kanjee Z, et al. Clinical Reasoning of a Generative Artificial Intelligence Model Compared With Physicians. JAMA Intern Med. 84(5):581–583. 2024. doi:10.1001/jamainternmed.2024.0295

3. Goh E, Gallo R, Hom J, et al. Large Language Model Influence on Diagnostic Reasoning: A Randomized Clinical Trial. JAMA Netw Open. 7(10):e2440969–e2440969. 2024. doi:10.1001/jamanetworkopen.2024.40969

4. McDuff D, Schaekermann M, Tu T, et al. Towards accurate differential diagnosis with large language models. Nature. 1–7. 2025. doi:10.1038/s41586-025-08869-4

5. Horvitz E. Principles of mixed-initiative user interfaces. Proceedings of the SIGCHI conference on Human Factors in Computing Systems (CHI ’99). Association for Computing Machinery, New York, NY, USA, 159–166. 1999. doi:10.1145/302979.303030

6. Amershi S, Weld D, Vorvoreanu M, et al. Guidelines for human-AI interaction. In Proceedings of the 2019 CHI Conference on Human Factors in Computing Systems (CHI ’19). Association for Computing Machinery, New York, NY, USA, Paper 3, 1–13. 2019. doi:10.1145/3290605.3300233

7. Wilder B, Horvitz E, Kamar E. Learning to complement humans. IJCAI’20: Proceedings of the Twenty-Ninth International Conference on International Joint Conferences on Artificial Intelligence. 212:1526–1533. 2020. doi:10.24963/ijcai.2020/212

8. Fogliato R, Chappidi S, Lungren M, et al. Who goes first? Influences of human-AI workflow on decision making in clinical imaging. Proceedings of the 2022 ACM Conference on Fairness, Accountability, and Transparency (FAccT ’22). Association for Computing Machinery, New York, NY, USA, 1362–1374. 2022. doi:10.1145/3531146.3533193

9. Ouyang L, Wu J, Jiang X, et al. Training language models to follow instructions with human feedback. Adv Neural Inf Process Syst. 35:27730–27744. 2022.

10. Sharma M, Tong M, Korbak T, et al. Towards understanding sycophancy in language models. ArXiv Prepr. Published online 2023. doi:10.48550/arXiv.2310.13548

11. Salecha A, Ireland ME, Subrahmanya S, Sedoc J, Ungar LH, Eichstaedt JC. Large language models display human-like social desirability biases in Big Five personality surveys. PNAS Nexus. 3(12):pgae533. 2024. doi:10.1093/pnasnexus/pgae533.

12. Sellen A, Horvitz E. The rise of the AI co-pilot: Lessons for design from aviation and beyond. Communications of the ACM. 67(7):18–23. 2024. doi:10.1145/3637865

13. Drosos I, Sarkar A, Toronto N. “ It makes you think”: Provocations Help Restore Critical Thinking to AI-Assisted Knowledge Work. *ArXiv Prepr*. Published online 2025. doi:10.48550/arXiv.2501.17247

14. Mozannar H, Satyanarayan A, Sontag D. Teaching Humans When to Defer to a Classifier via Exemplars. Proceedings of the AAAI Conference on Artificial Intelligence. 36, 5323–5331. 2022. doi:10.1609/aaai.v36i5.20469.

15. Mozannar H, Bansal G, Fourney A, Horvitz E. When to show a suggestion? Integrating human feedback in AI-assisted programming. Proceedings of the AAAI Conference on Artificial Intelligence, Vol. 38, No. 9, pp. 10137–10144. 2024. doi:10.1609/aaai.v38i9.28878

16. Shaikh O, Mozannar H, Bansal G, Fourney A, Horvitz E. Navigating Rifts in Human-LLM Grounding: Study and Benchmark. ACL 2025: Proceedings of 63rd Annual meeting of the Association for Computational Linguistics. 2025. doi:10.48550/arXiv.2503.13975

17. Tversky A, Kahneman D. Judgment under Uncertainty: Heuristics and Biases: Biases in judgments reveal some heuristics of thinking under uncertainty. Science. 185(4157):1124–1131. 1974. doi:10.1126/science.185.4157.11

18. Bansal G, Nushi B, Kamar E, Lasecki WS, Weld DS, Horvitz E. Beyond accuracy: The role of mental models in human-AI team performance. Proceedings of the AAAI Conference on Human Computation and Crowdsourcing, 7(1), 2–11. 2019. doi:10.1609/hcomp.v7i1.5285

19. Langlotz CP, Shortliffe EH. Adapting a consultation system to critique user plans. Int J Man-Mach Stud. 19(5):479–496. 1983. doi:10.1016/S0020-7373(83)80067-4

20. Miller PL. ATTENDING: Critiquing a physician’s management plan. IEEE Trans Pattern Anal Mach Intell. (5):449–461. 1983. doi:10.1109/TPAMI.1983.4767424

21. Bejnordi BE, Veta M, Van Diest PJ, et al. Diagnostic assessment of deep learning algorithms for detection of lymph node metastases in women with breast cancer. JAMA. 318(22):2199–2210. 2017. doi:10.1001/jama.2017.14585

22. Savage T, Wang J, Gallo R, et al. Large language model uncertainty proxies: discrimination and calibration for medical diagnosis and treatment. J Am Med Inform Assoc. 32(1):139–149. 2025. doi:10.1093/jamia/ocae254

23. Balachandran V, Chen J, Joshi N, et al. Eureka: Evaluating and understanding large foundation models. ArXiv Prepr. Published online 2024. doi:10.48550/arXiv.2409.10566

